# Optimizing the number of models included in outbreak forecasting ensembles

**DOI:** 10.1101/2024.01.05.24300909

**Authors:** Spencer J. Fox, Minsu Kim, Lauren Ancel Meyers, Nicholas G. Reich, Evan L. Ray

**Author notes:** Corresponding author: 1-706-542-9394, Spencer Fox 120 B.S. Miller Hall, Health Sciences Campus, 101 Buck Road, Athens, GA 30602.

## Abstract

Based on historical influenza and COVID-19 forecasts, we quantify the relationship between the number of models in an ensemble and its accuracy and introduce an ensemble approach that can outperform the current standard. Our results can assist collaborative forecasting efforts by identifying target participation rates and improving ensemble forecast performance.

## Text

Pioneered by the Centers for Disease Control and Prevention’s (CDC’s) 2013-2014 Influenza Season Challenge, real-time, collaborative forecast efforts have become the gold standard for generating and evaluating forecasts for infectious disease outbreaks (1,2). Individual component forecasts are aggregated into ensemble predictions that are the primary external communication provided by the organizing hubs and have consistently outperformed individual models (3–5). The current COVID-19 and influenza ensemble forecasts use the median across all eligible forecasts for each requested target, though other strategies that weight individual forecasts based on historical performance may further improve performance (6). To assist public health decision-makers considering target participation rates and the optimal design of ensemble forecast models, we retrospectively analyzed data from recent US-based collaborative outbreak forecast efforts to identify how the number of models included in an ensemble impacts performance.

We analyzed forecasts from five recent public collaborative forecast efforts including forecasts for influenza-like illness (ILI) from 2010-2017 (5), for COVID-19 reported cases, hospital admissions, and mortality from 2020-2023 (7), and for influenza hospital admissions from 2021-2023 (8). For each, we identified time periods with maximal model participation, *n* ∈ (1, …,*N*) created training and testing time periods, and obtained forecasts for individual, non-ensemble, models that produced at least 90% of all possible forecasts throughout those periods (Table S1). We created ensemble forecasts of size ***n* ∈ (1, …,*N*)**, where *n* is the number of individual models included in a given ensemble and *N* is the total number of available individual models, using three strategies: (1) randomly sampling combinations of *n* models (Random), (2) choosing the top individually performing *n* models from a training period (Individual rank), or (3) choosing the top performing ensemble of size *n* from a training period (Ensemble rank). We compared performance of all ensembles against a baseline model (Baseline) that produces forecasts based on historical seasonality for ILI (5) or flat forecasts for all other metrics (3), and an unweighted ensemble composed of all submitted models that is currently used in real-time as the gold standard forecast (Published ensemble). We summarized probabilistic ensemble forecast skill (Figure 1A) using the log score for ILI forecasts and the weighted interval score (WIS) for all others (9,10), and transformed scores as needed so that lower numbers indicate better performance (Figure 1B). Further methodological details are provided in the supplement.

**Figure 1:**
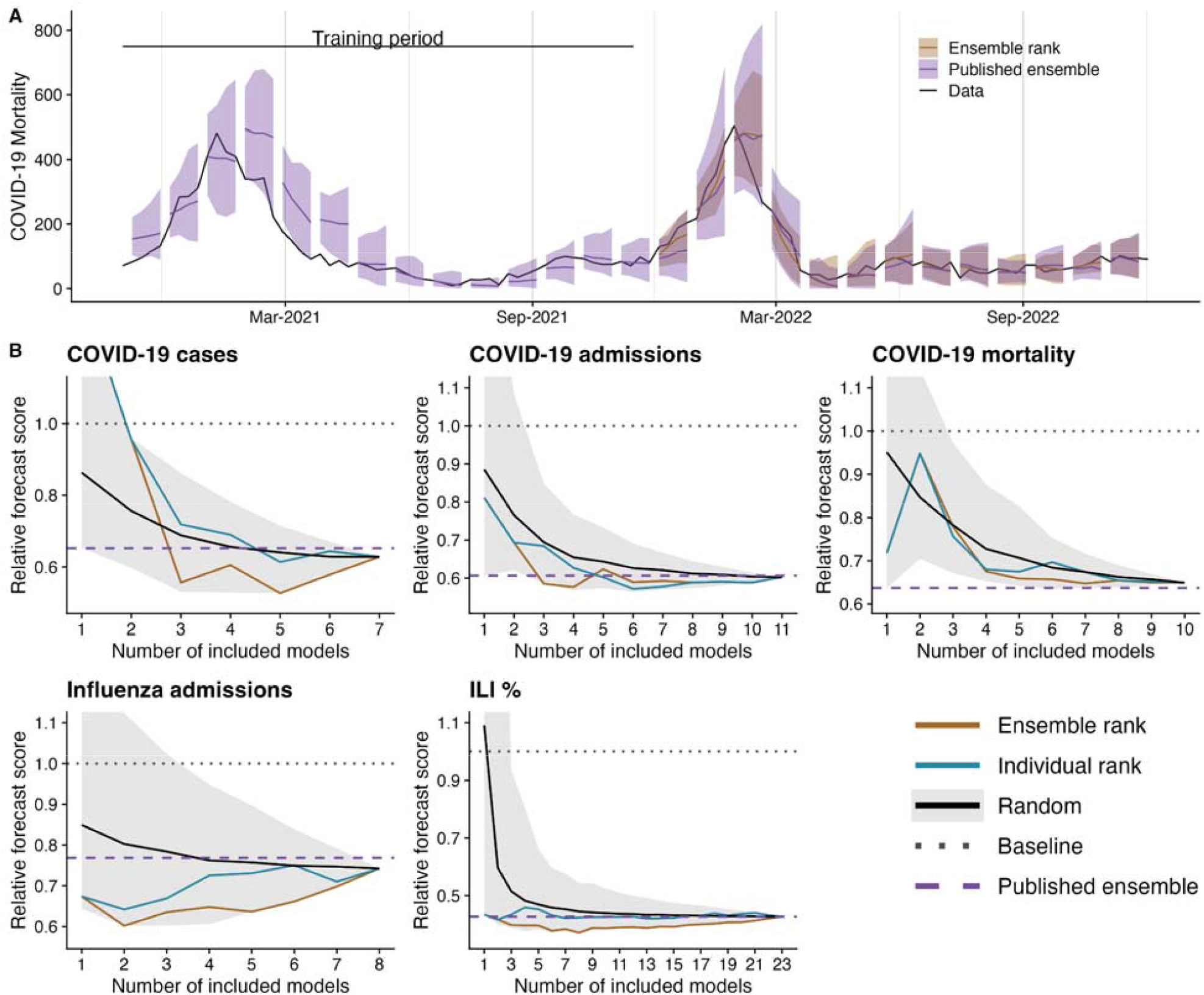
Forecast performance on recent influenza and COVID-19 collaborative forecast efforts comparing the number of models included in the ensemble and different ensemble methodologies. **(A)** Weekly COVID-19 mortality data for Massachusetts (black line) with forecasts from the published forecast that summarizes across all individual contributed forecasts (Published ensemble) and the best performing ensemble of size four from the training period (Ensemble rank). The line indicates the four-week point predictions and the shaded region indicates the 95% prediction interval for each ensemble. Ensemble rank forecasts require training and therefore are only shown during the testing period. **(B)** Summarized ensemble forecast scores from the collaborative forecast efforts for the weekly influenza-like illness (ILI) data provided by the CDC (ILI %), COVID-19 weekly case and mortality counts provided by JHU (COVID-19 cases and COVID-19 mortality), and COVID-19 and Influenza daily hospital admissions provided by HHS (COVID-19 admissions and Influenza admissions). Scores correspond to the average forecast performance during the respective testing periods across all dates, locations, and forecast horizons (Table S1). We plot the minimum (Grey region, lower), maximum (Grey region, upper), and mean (Solid black line) scores of random ensemble combinations of a given size (Random), and the trained ensembles composed of the top *n* individual performing models from the training period (Individual rank) or the best performing ensemble of size *n* from the training period (Ensemble rank). All scores are standardized by the baseline forecast model for that metric (horizontal dotted line), and the horizontal dashed line corresponds to the Published ensemble that is the unweighted ensemble across all models that submitted for a specific date and forecast target and is used as the gold-standard forecast prediction. Relative scores less than 1 indicate better accuracy than the Baseline. On average across the testing phase, the Published ensemble included 15 models for COVID-19 cases, 17 models for COVID-19 admissions, 19 models for COVID-19 deaths, 21 models for influenza admissions, and 23 models for ILI.

When using random sampling for choosing component ensemble models, we found that for all forecasting exercises, including more models yielded better average forecast performance and all ensembles outperformed the Baseline model after the inclusion of at least four models (Figure 1B). Increasing the ensemble size above four models only slightly improved the average forecast performance, but substantially decreased the variability of performance across randomly assembled models. For example, for influenza hospital admission forecasts, increasing the number of models in the ensemble from four to seven improved the average ensemble performance by 2%, but reduced the interquartile range across possible ensembles by 56.5%. In essence, increasing the ensemble size increases the likelihood that a randomly chosen ensemble performs well.

Ensemble creation based on the individual rank order performance from the training period gave mixed forecast results, while creation based on historical ensemble performance consistently selected high-performing ensembles that prospectively beat or matched the Published ensemble (Figure 1B, Table 1). For the Ensemble rank method, performance generally plateaued or declined when more than four models were included for both the testing (Figure 1) and training period (Figure S1). The Ensemble rank of size four had relative forecast performance against the Published ensemble of 0.94 and 0.84 for ILI and influenza hospital admissions, respectively, and 0.93, 0.95, and 1.06 for COVID-19 cases, hospital admissions, and mortality, respectively, where values less than 1 indicate performance improvements. While the Ensemble rank model did not always match the prediction interval coverage of the Published ensemble (Figure S2), its average rank for individual prediction tasks was always better than that of the Published ensemble (Figure S3-S7). We found that relative forecast performance is consistent when viewed across the different locations, dates, and targets (Figure S8-S16).

**Table 1:** Forecast performance of ensembles of size four relative to the Published ensemble forecast model for each of the respective collaborative forecast efforts, where values less than 1 indicate improved forecast performance. On average across the testing phase, the Published ensemble included 15 models for COVID-19 cases, 17 models for COVID-19 admissions, 19 models for COVID-19 deaths, 21 models for influenza admissions, and 23 models for ILI.

|  | Random model (range) | Individual rank | Ensemble rank |
| --- | --- | --- | --- |
| COVID-19 Cases | 1.01 (0.81-1.2) | 1.06 | 0.93 |
| COVID-19 Admits | 1.08 (0.94-1.27) | 1.03 | 0.95 |
| COVID-19 Deaths | 1.14 (1.02-1.37) | 1.07 | 1.06 |
| ILI % | 1.13 (0.89-1.9) | 1.08 | 0.94 |
| Influenza Admits | 0.99 (0.79-1.23) | 0.94 | 0.84 |

While our results are constrained by a limited number of diseases, forecasting exercises, and models to draw upon, they have several implications for future collaborative forecast efforts: (1) increasing participation in collaborative forecast hubs increases model diversity, improves the average forecast performance, and decreases variability between possible ensemble combinations, (2) while optimizing ensemble forecasts based on historical performance does not guarantee optimal future performance, data-driven selection of models can improve forecast performance compared to unweighted ensembles, and (3) evaluating ensemble rather than individual performance selects for complementarity in forecasts and consistently improved forecast performance. As public health officials and researchers look to expand collaborative forecast efforts and as funding agencies allocate budgets across methodological and applied forecast efforts, our results can be used to identify target participation rates, guide the interpretation and communication of ensemble forecasts, and improve forecast performance.

## Data Availability

All forecasts and ground truth data used in the analysis are publicly available in their specific forecast repositories. Code that gathers the data from the individual competitions and replicates the analysis presented in this manuscript can be found at https://github.com/sjfox/ensemble-size.

## Acknowledgments

The authors acknowledge the helpful comments from the members of the CSTE, CDC, and MIDAS forecasting working groups as well as the Scenario Modeling Hub. The authors also acknowledge the Texas Advanced Computing Center (TACC) at The University of Texas at Austin for providing HPC resources that have contributed to the research results reported within this paper. URL: http://www.tacc.utexas.edu. SJF and LAM were supported by the Council for State and Territorial Epidemiologists (NU38OT000297) and the CDC (75D30122C14776). MK, ELR, and NGR were supported by the National Institutes of General Medical Sciences (R35GM119582) and the US CDC (1U01IP001122). The content is solely the responsibility of the authors and does not necessarily represent the official views of CSTE, CDC, NIGMS, or the National Institutes of Health.

## Biographical Sketch

Dr. Spencer J. Fox is an Assistant Professor at the University of Georgia in the Department of Epidemiology & Biostatistics and the Institute of Bioinformatics. His research interests include statistical modeling of emerging infectious diseases and outbreak forecasting.

## Conflicts of Interest

The authors declare no conflicts of interest.

## Notes

### Competing Interest Statement

The authors have declared no competing interest.

