## Supplemental Information for "Optimizing the number of models included in outbreak forecasting ensembles"

Spencer J. Fox^1,2,3^

Minsu Kim^4^

Nicholas G. Reich^4^

Evan L. Ray^4^

#### Affiliations:

^1^ - Department of Epidemiology & Biostatistics, University of Georgia

^2^ - Institute of Bioinformatics, University of Georgia

^3^ - Center for Ecology of Infectious Diseases, University of Georgia

^4^ - Department of Biostatistics and Epidemiology, University of Massachusetts Amherst

^5^ - Department of Integrative Biology, University of Texas at Austin

^6^ - Department of Statistics and Data Science, University of Texas at Austin

^7^ - Department of Population Health, Dell Medical School

### Supplemental methodology

#### Methodological overview

The following sections describe our approach to (1) choosing the models and time periods to include for each forecast effort; (2) constructing ensemble forecasts for the selected retrospective time periods; (3) creating and scoring ensemble forecasts for random subsets of available individual models; and (4) constructing the individual rank and ensemble rank ensembles with selected component models.

#### Collaborative forecast competitions investigated

To ensure our ensemble analysis results were consistent and robust, we carried out the ensemble analysis on all United States-based collaborative forecast efforts with publicly available submission formats (i.e. those that organized submissions using a hub format on Github). These include the coordinated efforts to forecast influenza-like illness percent over seven influenza seasons (1), influenza hospitalizations beginning in 2021 (2), and COVID-19 case counts, hospital admissions, and mortality beginning in 2020 (3–5).

#### Model and time period selection

To carry out the analysis we selected time periods for each collaborative forecast effort that had the maximum number of individual component forecast models with submissions for at least 90% of all possible forecasts. We included the baseline forecast models and we excluded all ensembles of forecasts from other models that contributed to each competition as that follows the inclusion criteria for the current Published ensemble model. We also included the Published ensemble for each competition to be used as a reference for comparison, but these models were not incorporated as possible members of the ensembles that we created. We split up the time periods into training and testing periods, ensuring that each period for each metric had at least a period of epidemic growth and decline. The final time periods, included models, and specified baseline models can be found in Table S1.

**Table S1**: Selected time periods and models for each collaborative forecast effort. Baseline models for each metric are specified in bold.

| Disease | Metric | Training period | Testing period | Models included |
| --- | --- | --- | --- | --- |
| COVID-19 | Cases | Nov 02, 2020 - Nov 08, 2021 | Nov 15, 2021 - July 25, 2022 | 1. BPagano-RtDriven 2. CovidAnalytics-DELPHI 3. **COVIDhub-baseline** 4. CU-select 5. JHUAPL-Bucky 6. RobertWalraven-ESG 7. USC-SI_kJalpha |
|  | Hospital Admissions | Feb 07, 2022 - May 23, 2022 | May 30, 2022 - Nov 07, 2022 | 1. BPagano-RtDriven 2. **COVIDhub-baseline** 3. CMU-TimeSeries 4. CU-select 5. CUB_PopCouncil-SLSTM 6. GT-DeepCOVID 7. Karlen-pypm 8. MOBS-GLEAM_COVID 9. MUNI-ARIMA 10. PSI-DICE 11. USC-SI_kJalpha |
|  | Deaths | Nov 02, 2020 - Nov 08, 2021 | Nov 15, 2021 - Nov 07, 2022 | 1. BPagano-RtDriven 2. **COVIDhub-baseline** 3. CU-select 4. GT-DeepCOVID 5. Karlen-pypm 6. MOBS-GLEAM_COVID 7. PSI-DRAFT 8. RobertWalraven-ESG 9. USC-SI_kJalpha 10. UCSD_NEU-DeepGLEAM |
| Influenza | Hospital Admissions | Jan 10, 2022 - Jun 20, 2022 | Oct 17, 2022 - April 03, 2023 | 1. CMU-TimeSeries 2. **Flusight-baseline** 3. GT-FluFNP 4. MOBS-GLEAM_FLUH 5. PSI-DICE 6. SGroup-RandomForest 7. SigSci-CREG 8. SigSci-TSENS |
|  | Influenza-like Illness (%) | Flu seasons: '2010/2011', '2011/2012', '2012/2013', '2013/2014' | Flu seasons:  '2014/2015', '2015/2016', '2016/2017', | 1. CU_EAKFC_SEIRS 2. CU_EAKFC_SIRS 3. CU_EKF_SEIRS 4. CU_EKF_SIRS 5. CU_RHF_SEIRS 6. CU_RHF_SIRS 7. CUBMA 8. Delphi_BasisRegression 9. Delphi_EmpiricalFutures 10. Delphi_ExtendedDeltaDensity 11. Delphi_MarkovianDeltaDensity 12. FluOutlook_Mech 13. FluOutlook_MechAug 14. FluX_ARLR 15. FluX_LSTM 16. LANL_DBMplus 17. Protea_Kudu 18. Protea_Springbok 19. ReichLab_kcde_backfill_none 20. **ReichLab_kde** 21. ReichLab_sarima_seasonal_difference_FALSE 22. ReichLab_sarima_seasonal_difference_TRUE 23. UA_EpiCos |

#### Ensemble creation, forecasting, and scoring

For all but the multiyear ILI % competition, we created ensembles for all possible combinations of individual models of a specified size, *n*, for all values of n from 1 to N where N is the total number of models included for that forecasting exercise (Table S1). This yielded 127, 2047, 1023, and 255 total ensembles for the COVID-19 case counts, COVID-19 hospital admissions, COVID-19 mortality counts, and influenza hospital admissions respectively. For these competitions, we followed the methodology used to create real-time, published forecast ensembles in (4) and created median ensemble models for all forecast dates and targets for the specified individual component models using the hubEnsembles R package (6). As not every model submitted forecasts for every date and horizon, some ensemble forecasts of a specified size, *n*, had fewer than *n* component models. We did not exclude these from our analysis, because it was a rare occurrence and it replicates the real world scenario where some models will miss some submissions. We scored all forecasts for all ensemble models following the scoring methods from (4) using the methodologies made available in the covidHubUtils R package (7). We focused our analysis on the weighted interval score (WIS), which captures overall forecast performance, and the prediction interval coverage, which estimates the calibration of forecast uncertainty (7–9). Following the methods in (4), we analyzed the average WIS and prediction interval coverage for only forecasts of the states and territories, as national-level forecasts can skew forecast performance estimates. For these forecasting exercises, the Published ensemble was the ensemble produced by the hub using all available models (including those that did not meet our eligibility criteria); thus, the Published ensemble generally included more models than the ensembles that we considered.

For the multiyear ILI % analysis, we followed a different creation and scoring methodology due to the different forecast format and the number of individual component models. For this competition, teams were asked to submit 100% of all forecast targets and dates retrospectively, so we included all 23 individual component models that were successfully submitted on GitHub (10). Given the 23 models, there would be 8,388,607 total possible ensemble combinations, which was computationally impractical to run, score, and post-process. For ensembles of size *n* that had fewer than 1,000 possible combinations, we ran all of those combinations, but for those that had more than 1,000 possible combinations, we randomly selected 1,000 ensembles from the options. In total, we included 18,553 total ensemble combinations in the analysis and we used the largest ensemble of size 23 as the Published ensemble. We produced ensemble forecasts for all targets and dates using the ensemble methodology used in (11). As these forecasts were submitted in a bin probability format rather than an interval format, we followed the methodology described in (11) to create a linear pool ensemble model that assumes equal weights across all included component models for all forecast dates and targets. To do so we utilized publicly available code provided in (12). We followed the scoring methodology of (1) and used the FluSight R package functions to score all ensemble forecasts (13). Following the methodology described in (11) we summarized the individual forecast scores for every date and target as
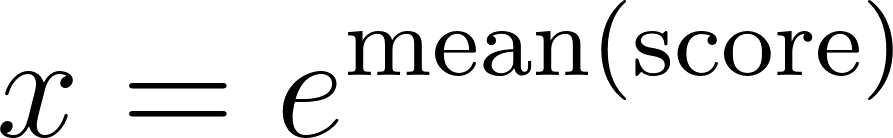
, where
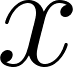
 is the resulting summary forecast score. We took
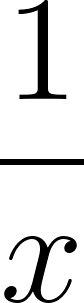
 as the final measure of forecast skill, so that smaller values indicated better forecast performance consistent with the interpretation of WIS.

#### Ensemble methodology

In our main analysis where we only investigate ensemble forecast performance during the testing period, we present results from three different ensemble methodologies

1. A random sampling methodology (Random) that presents the forecast scores for ensembles of size *n* across all created ensembles of that size in the testing period. Since we create all possible combinations of ensembles for all but the multiyear ILI % analysis, presenting these results is equivalent to presenting range and average results if one were to randomly combine models to achieve a specified ensemble size. For the multiyear ILI % analysis, in settings where the number of created ensembles was capped at 1,000 it presents the range and mean of scores for the ensembles of randomly selected individual models.
2. A component model selection scheme that relies on the individual rank of the component models from the training period (Individual rank). To create an ensemble of size *n*, we choose the top *n* individually performing models from the training period to use as the members of the ensemble in the testing period. Results from these ensembles are only presented in the testing period.
3. A component model selection scheme that relies on the ensemble rank of the investigated ensemble models from the training period (Ensemble rank). For this model, we create an ensemble of a specified size *n* by identifying the ensemble of size *n* that had the best forecast performance in the training period. An ensemble using those same component models was then used to generate forecasts for the testing period. For the multiyear ILI % analysis, not all ensemble combinations were created and analyzed in the training period (as described in the previous section), so we limited our ensemble choice to only those that were analyzed from the random sample created. This means we may not have chosen the best performing ensemble from the training period across all possible options.

#### Data and code availability

All forecasts and ground truth data used in the analysis are publicly available in their specific forecast repositories. Code that gathers the data from the individual competitions and replicates the analysis presented in this manuscript can be found at https://github.com/sjfox/ensemble-size.

### Supplemental figures and tables

#### Training period performance results

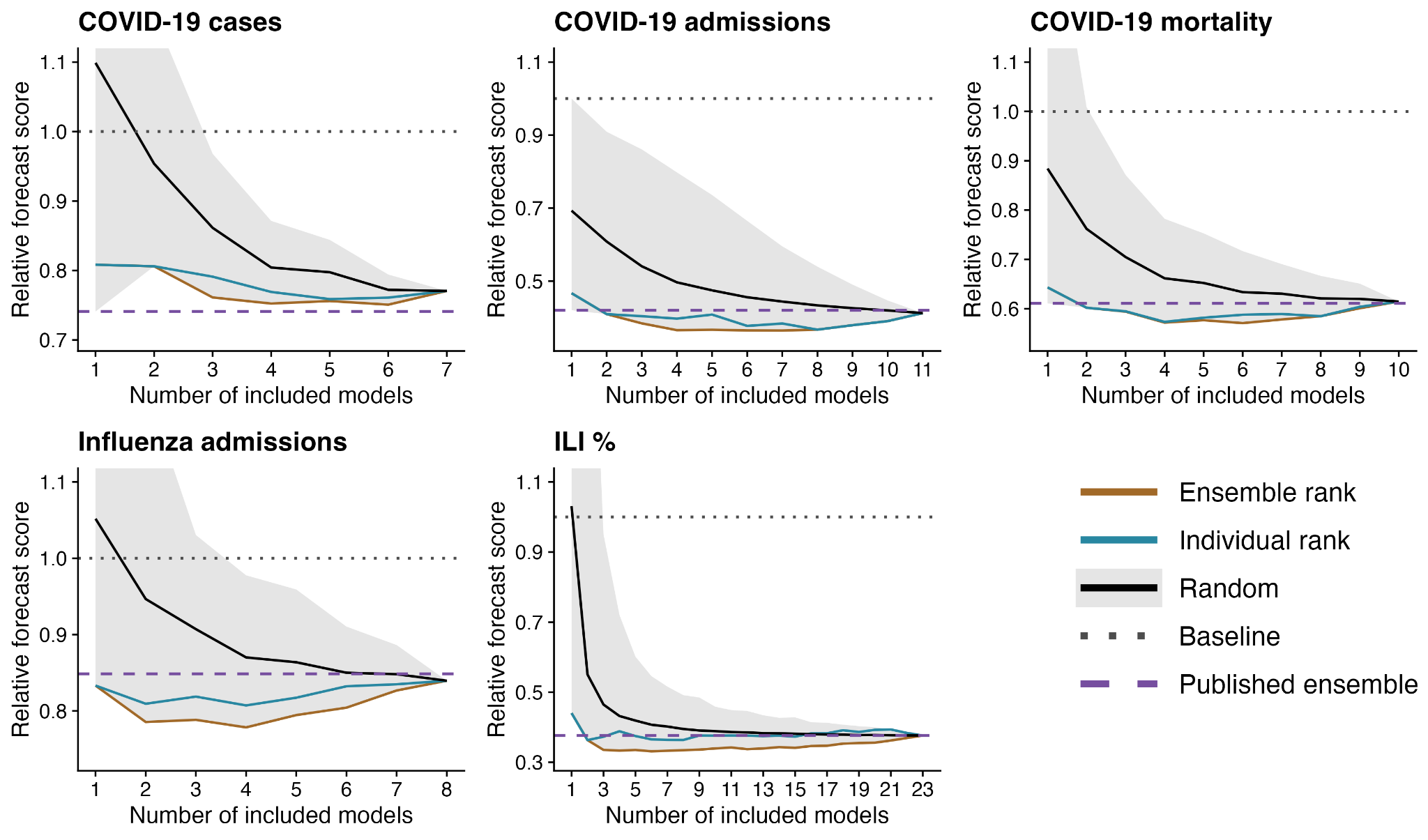

**Figure S1: Forecast performance on the training period for recent influenza and COVID-19 collaborative forecast efforts comparing the number of models included in the ensemble and different ensemble methodologies.** Summarized ensemble forecast scores from the collaborative forecast efforts for the weekly influenza-like illness (ILI) data provided by the CDC (ILI %), COVID-19 weekly case and mortality counts provided by JHU (COVID-19 cases and COVID-19 mortality), and COVID-19 and Influenza daily hospital admissions provided by HHS (COVID-19 admissions and Influenza admissions). Scores correspond to the average forecast performance during the respective training periods across all dates, locations, and forecast horizons (Table S1). We plot the minimum (Grey region, lower), maximum (Grey region, upper), and mean (Solid black line) scores of random ensemble combinations of a given size (Random), and the ensembles composed of the top n individual performing models from the training period (Individual rank) or the best performing ensemble of size n from the training period (Ensemble rank). These are included as a comparison with their ensemble performance in the testing period in the main manuscript. All scores are standardized by the baseline forecast model for that metric (horizontal dotted line), and the horizontal dashed line corresponds to the Published ensemble that is the unweighted ensemble across all models that submitted for a specific date and forecast target and is used as the gold-standard forecast prediction. Relative scores less than 1 indicate better accuracy than the Baseline.

#### Prediction interval coverage results

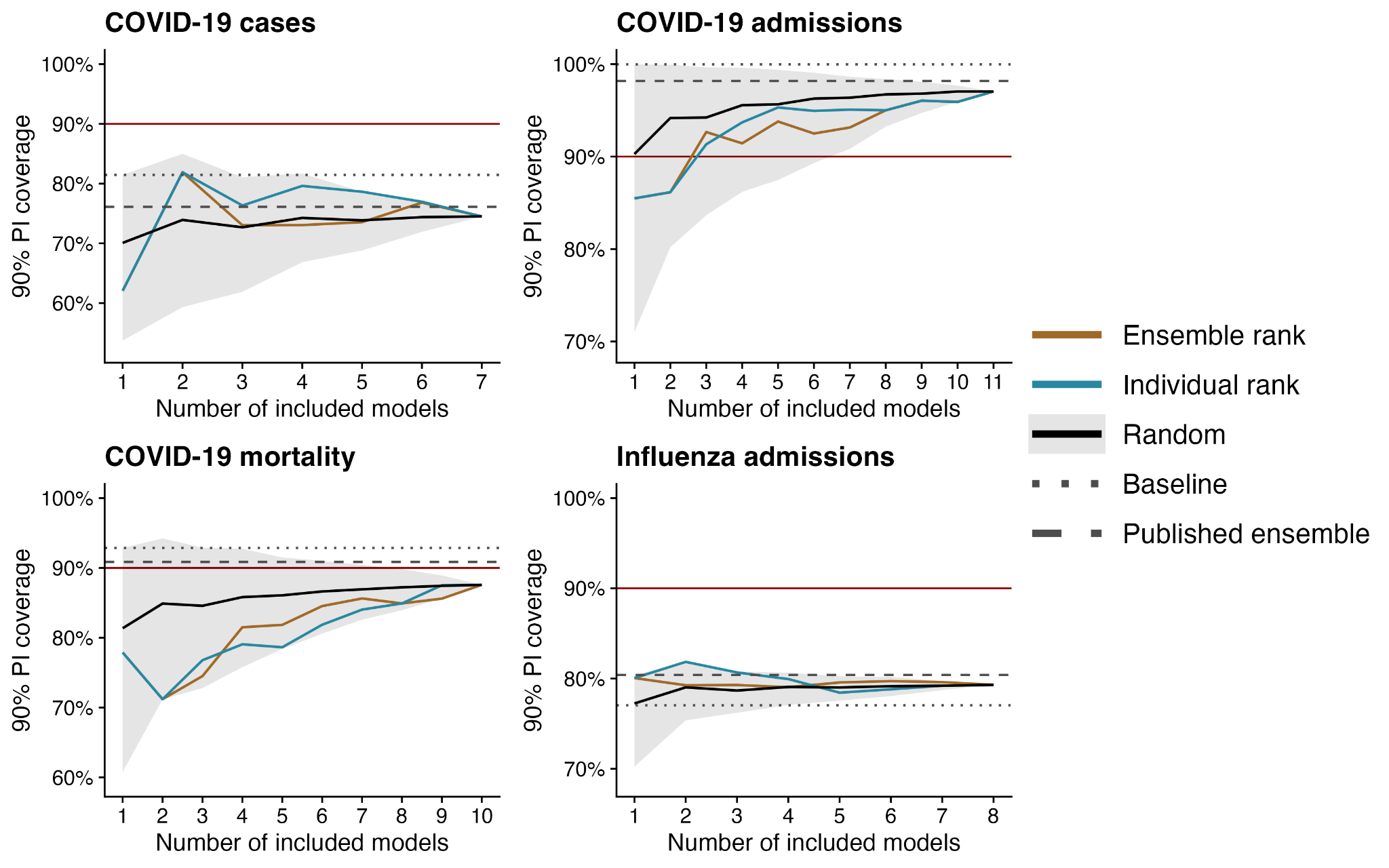

**Figure S2: Forecast prediction interval coverage (PIC) comparison for ensembles of varying ensemble size and component selection strategy on recent influenza and COVID-19 collaborative forecast efforts.** Summarized 90% PIC from the recent COVID-19 and influenza forecasting efforts during the respective testing periods across all dates, locations, and forecast horizons (Table S1). Well calibrated models are expected to have PIC near 90% (Red horizontal line). We plot the minimum (Grey ribbon, lower), maximum (Grey ribbon, upper), and mean (Solid black line) of random ensemble combinations of a given size (Random), and the trained ensembles composed of the top *n* individual performing models from the training period (Individual rank) or the best performing ensemble of size *n* from the training period (Ensemble rank). We plot the coverage rates from these models alongside the baseline forecast model that makes flat line predictions (horizontal dotted line), and the horizontal dashed line corresponds to the ensemble published in real-time (Published ensemble) that is the unweighted ensemble across all models that submitted for a specific date and forecast target and is used as the gold-standard forecast prediction.

#### Model score rankings by metric

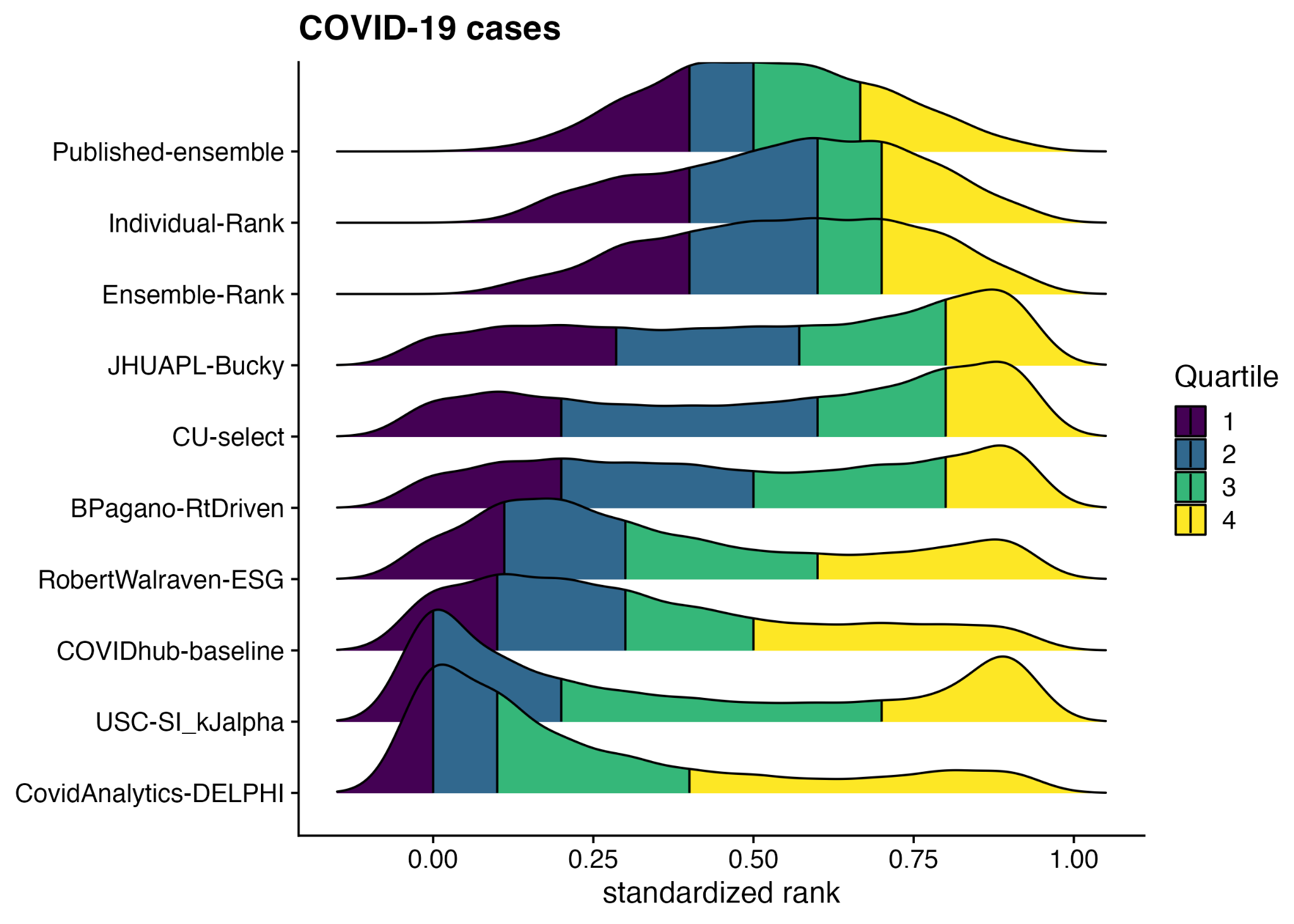

**Figure S3: Distribution of standardized weighted interval score (WIS) rank for forecasts of COVID-19 case counts across every forecasted date, location, and target in the testing period of the analysis.** A value of 0 indicates the model had the worst WIS for that particular location, target, and date while a value of 1 indicates that the model had the best WIS. Any density below zero comes from the smoothing of the density plot and should be interpreted as a value of zero. The quartiles of each model’s distribution of standardized ranks are shown in different colors: yellow indicates the top quarter of the distribution and purple indicates the bottom quarter of the distribution. The models are ordered by the 25th percentile distribution, with better forecasting models closer to the top. Results for Ensemble and Individual rank models of size four shown.

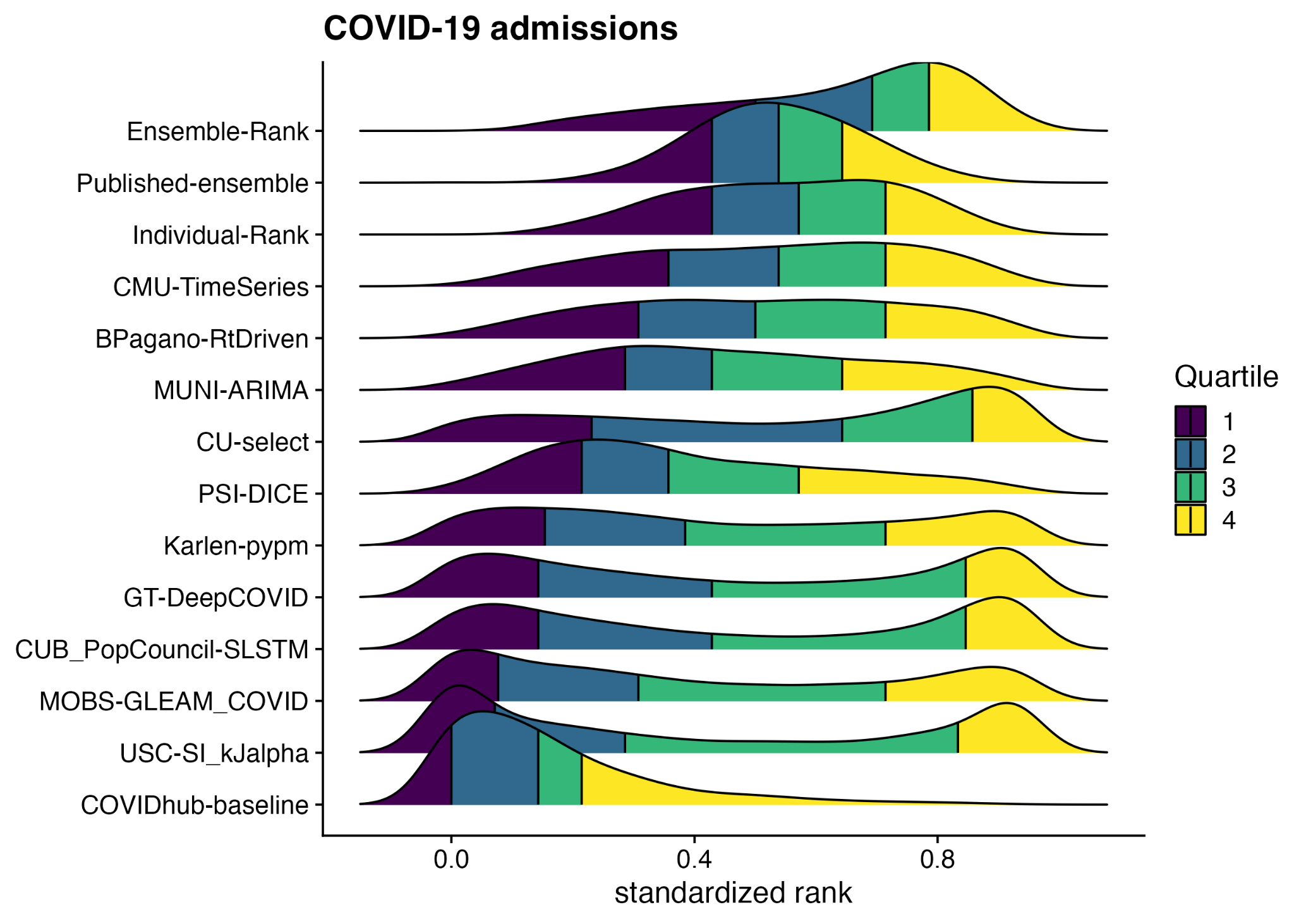

**Figure S4: Distribution of standardized weighted interval score (WIS) rank for forecasts of COVID-19 hospital admissions across every forecasted date, location, and target in the testing period of the analysis.** A value of 0 indicates the model had the worst WIS for that particular location, target, and date while a value of 1 indicates that the model had the best WIS. Any density below zero comes from the smoothing of the density plot and should be interpreted as a value of zero. The quartiles of each model’s distribution of standardized ranks are shown in different colors: yellow indicates the top quarter of the distribution and purple indicates the bottom quarter of the distribution. The models are ordered by the 25th percentile distribution, with better forecasting models closer to the top. Results for Ensemble and Individual rank models of size four shown.

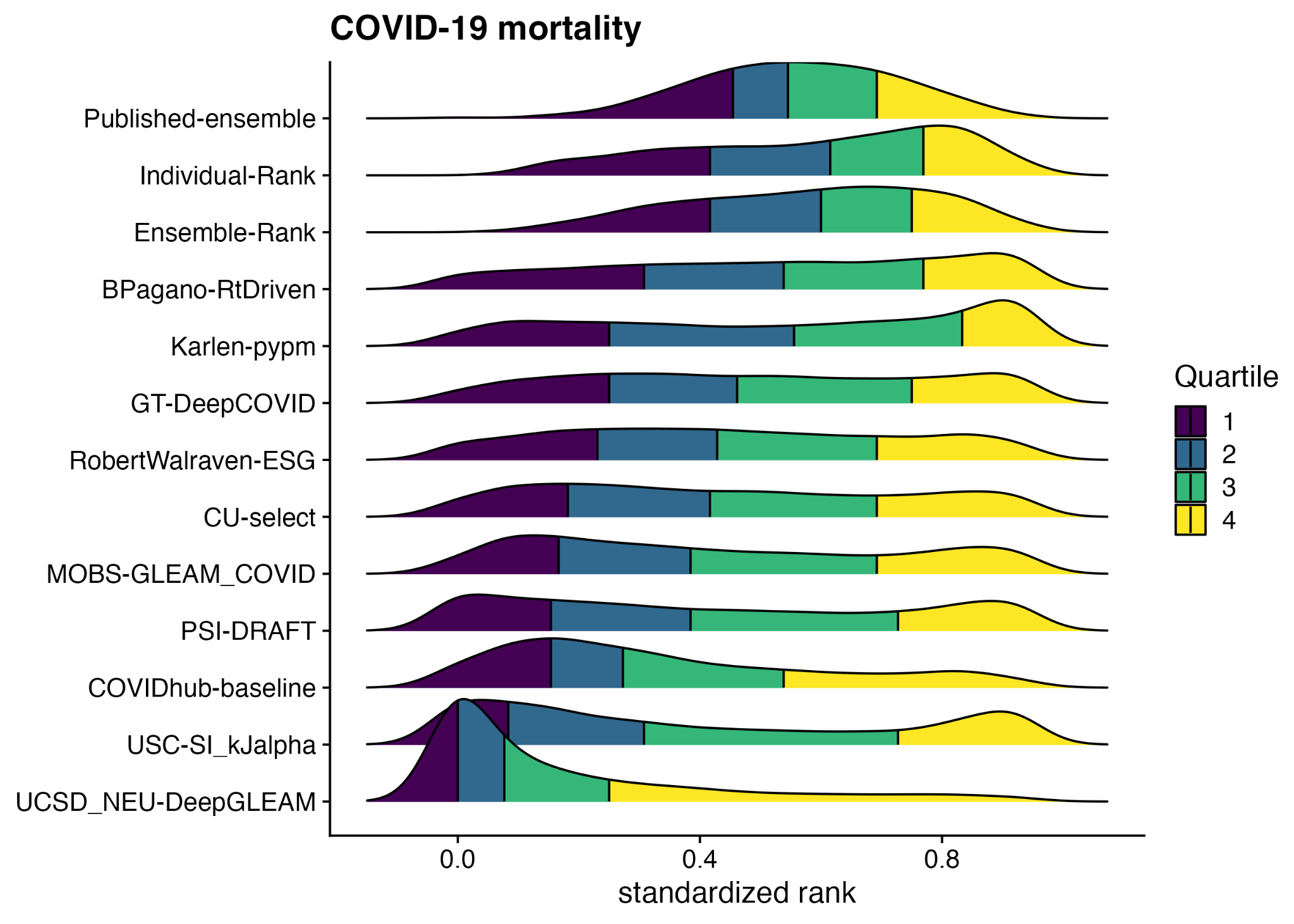

**Figure S5: Distribution of standardized weighted interval score (WIS) rank for forecasts of COVID-19 mortality across every forecasted date, location, and target in the testing period of the analysis.** A value of 0 indicates the model had the worst WIS for that particular location, target, and date while a value of 1 indicates that the model had the best WIS. Any density below zero comes from the smoothing of the density plot and should be interpreted as a value of zero. The quartiles of each model’s distribution of standardized ranks are shown in different colors: yellow indicates the top quarter of the distribution and purple indicates the bottom quarter of the distribution. The models are ordered by the 25th percentile distribution, with better forecasting models closer to the top. Results for Ensemble and Individual rank models of size four shown.

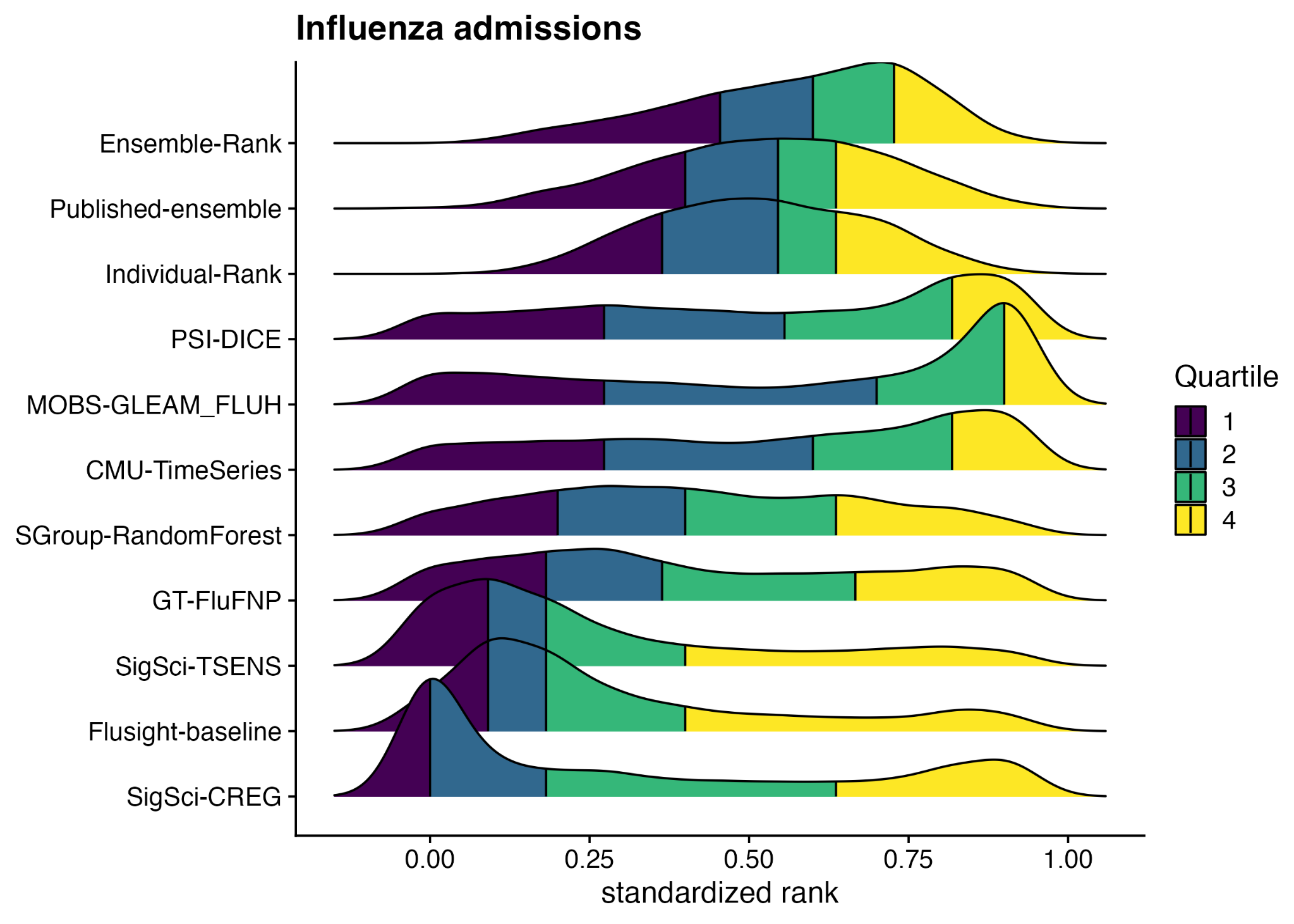

**Figure S6: Distribution of standardized weighted interval score (WIS) rank for forecasts of influenza hospital admissions across every forecasted date, location, and target in the testing period of the analysis.** A value of 0 indicates the model had the worst WIS for that particular location, target, and date while a value of 1 indicates that the model had the best WIS. Any density below zero comes from the smoothing of the density plot and should be interpreted as a value of zero. The quartiles of each model’s distribution of standardized ranks are shown in different colors: yellow indicates the top quarter of the distribution and purple indicates the bottom quarter of the distribution. The models are ordered by the 25th percentile distribution, with better forecasting models closer to the top. Results for Ensemble and Individual rank models of size four shown.

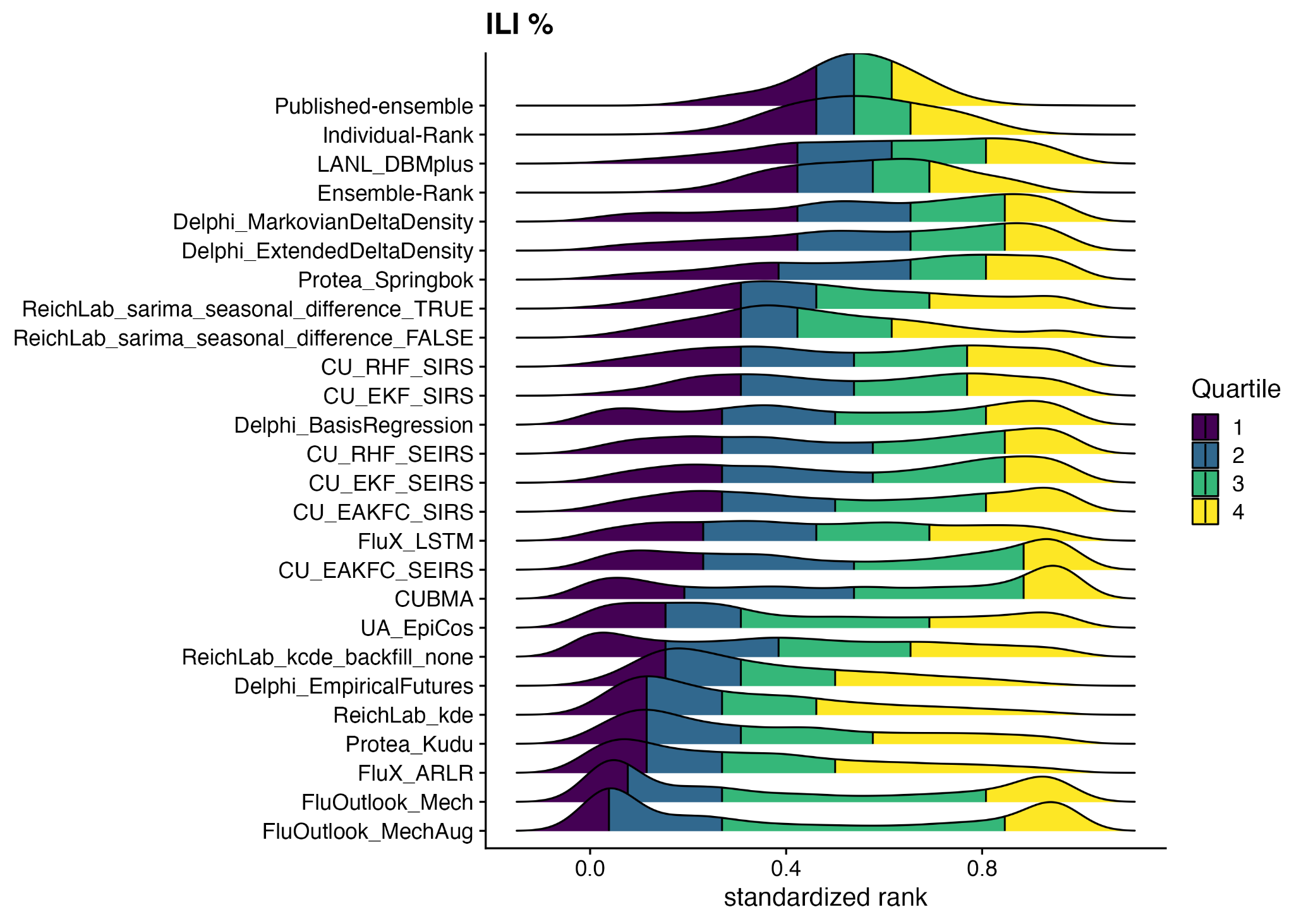

#### **Figure S7: Distribution of the standardized forecast score rank for forecasts of influenza-like illness (ILI %) across every forecasted date, location, and target in the testing period of the analysis.** A value of 0 indicates the model had the worst forecast score for that particular location, target, and date while a value of 1 indicates that the model had the best forecast score. Any density below zero comes from the smoothing of the density plot and should be interpreted as a value of zero. The quartiles of each model’s distribution of standardized ranks are shown in different colors: yellow indicates the top quarter of the distribution and purple indicates the bottom quarter of the distribution. The models are ordered by the 25th percentile distribution, with better forecasting models closer to the top. Results for Ensemble and Individual rank models of size four shown.

#### Date-specific epidemiological trends and forecast performance

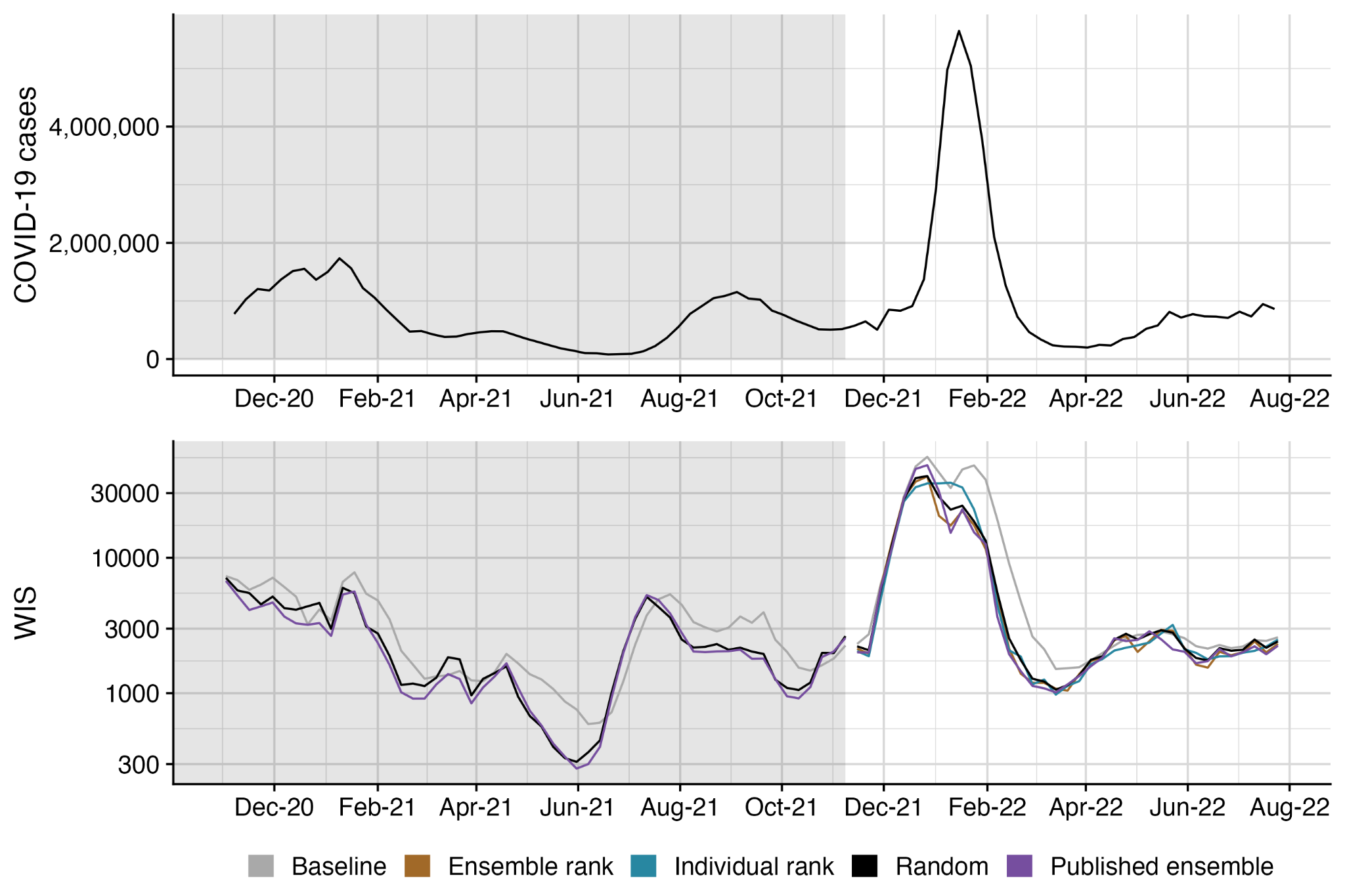

**Figure S8: COVID-19 case counts and model performance by date.** (Top) COVID-19 case counts nationally for the United States. (Bottom) Average weighted interval score (WIS) for each model and forecast date across all locations and targets from analysis. Lower scores indicate better forecast performance. Performance is visualized for ensembles of size four for the Ensemble rank, Individual rank, and Random ensembles, and only the mean performance is shown for the Random ensemble. Grey shaded region indicates the training period for ensemble rank and individual rank trained ensembles.

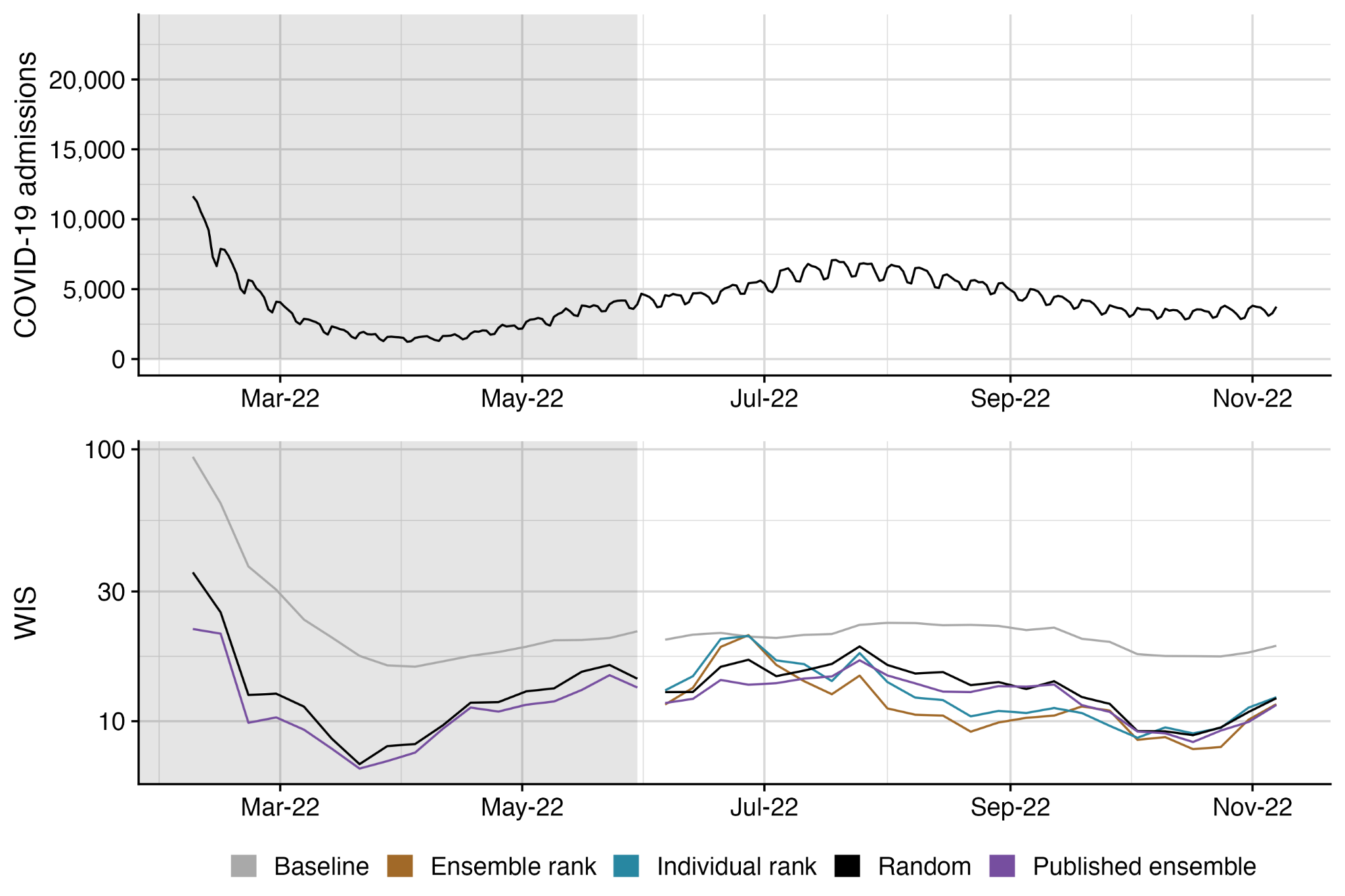
**Figure S9: COVID-19 hospital admissions and model performance by date.** (Top) COVID-19 hospital admissions nationally for the United States. (Bottom) Average weighted interval score (WIS) for each model and forecast date across all locations and targets from analysis. Lower scores indicate better forecast performance. Performance is visualized for ensembles of size four for the Ensemble rank, Individual rank, and Random ensembles, and only the mean performance is shown for the Random ensemble. Grey shaded region indicates the training period for ensemble rank and individual rank trained ensembles.

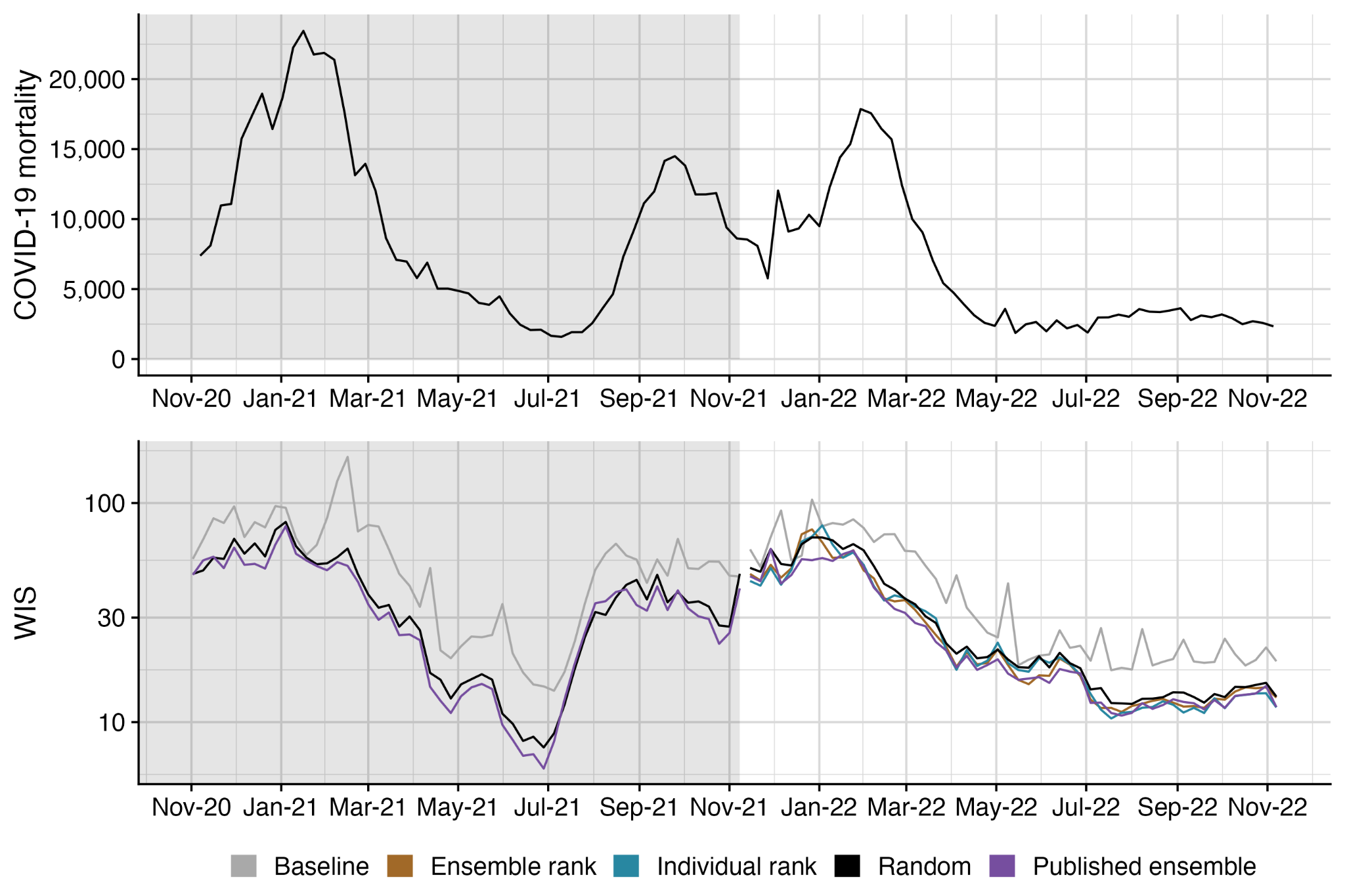
**Figure S10: COVID-19 mortality and model performance by date.** (Top) COVID-19 mortality nationally for the United States. (Bottom) Average weighted interval score (WIS) for each model and forecast date across all locations and targets from analysis. Lower scores indicate better forecast performance. Performance is visualized for ensembles of size four for the Ensemble rank, Individual rank, and Random ensembles, and only the mean performance is shown for the Random ensemble. Grey shaded region indicates the training period for ensemble rank and individual rank trained ensembles.

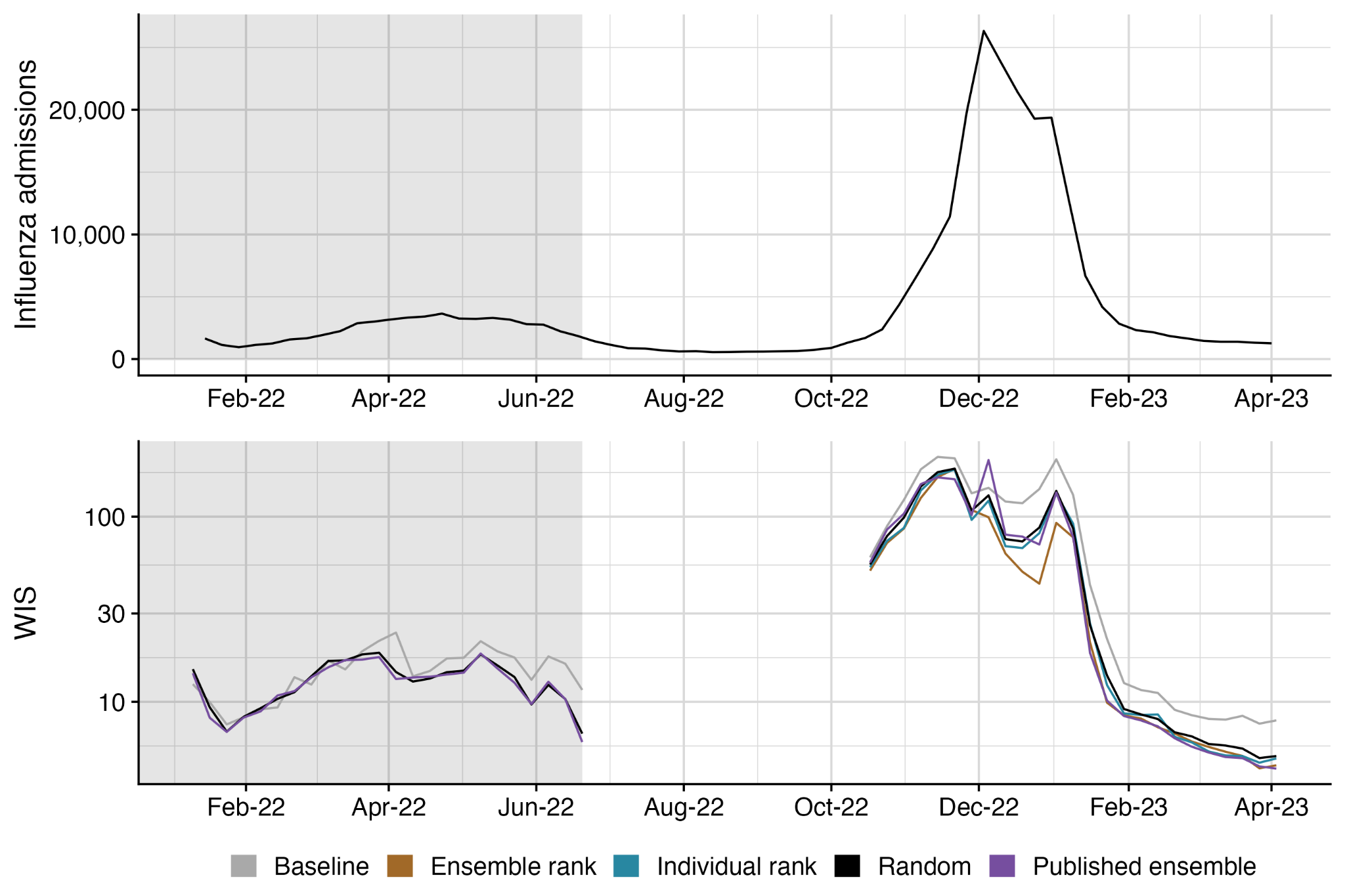

**Figure S11: Influenza hospital admissions and model performance by date.** (Top) Influenza hospital admissions nationally for the United States. (Bottom) Average weighted interval score (WIS) for each model and forecast date across all locations and targets from analysis. Lower scores indicate better forecast performance. Grey shaded region indicates the training period for ensemble rank and individual rank trained ensembles. Performance is visualized for ensembles of size four for the Ensemble rank, Individual rank, and Random ensembles, and only the mean performance is shown for the Random ensemble. Performance is not measured during the summer when the collaborative forecast efforts were paused.

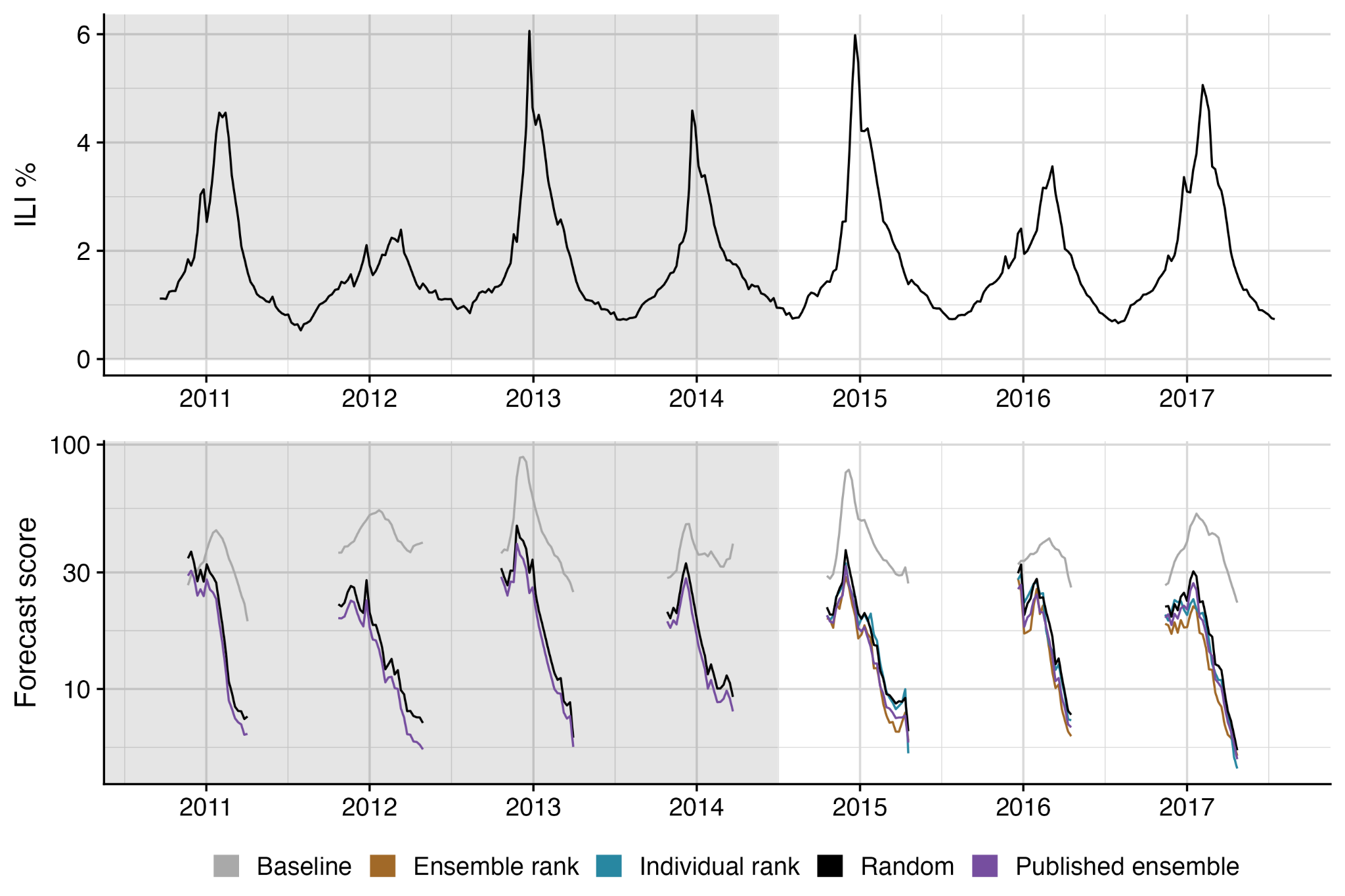

**Figure S12: Influenza-like illness (ILI %) and model performance by date.** (Top) ILI % nationally for the United States. (Bottom) Average forecast score for each model and forecast date across all locations and targets from analysis. Lower scores indicate better forecast performance. Grey shaded region indicates the training period for ensemble rank and individual rank trained ensembles. Performance is visualized for ensembles of size four for the Ensemble rank, Individual rank, and Random ensembles, and only the mean performance is shown for the Random ensemble. Performance is not measured during the summer months when the collaborative forecast efforts were paused.

#### Target-specific forecast performance

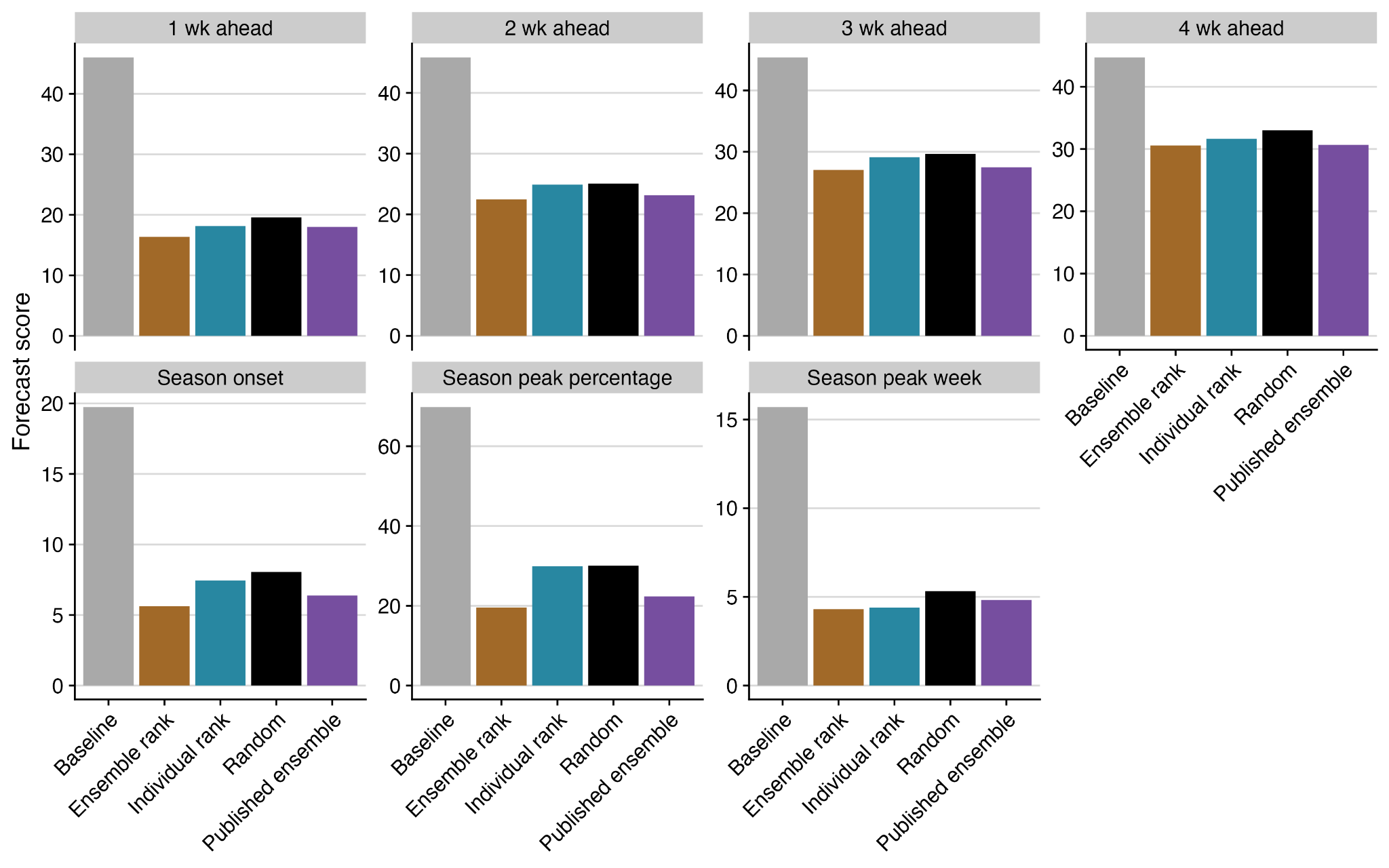

**Figure S13: Influenza-like illness (ILI %) model performance for each forecasted target.** Average forecast score for each model and each target (facets) across all forecast dates and locations from the testing period of the analysis. Lower scores indicate better forecast performance. Performance is visualized for ensembles of size four for the Ensemble rank, Individual rank, and Random ensembles, and only the mean performance is shown for the Random ensemble.

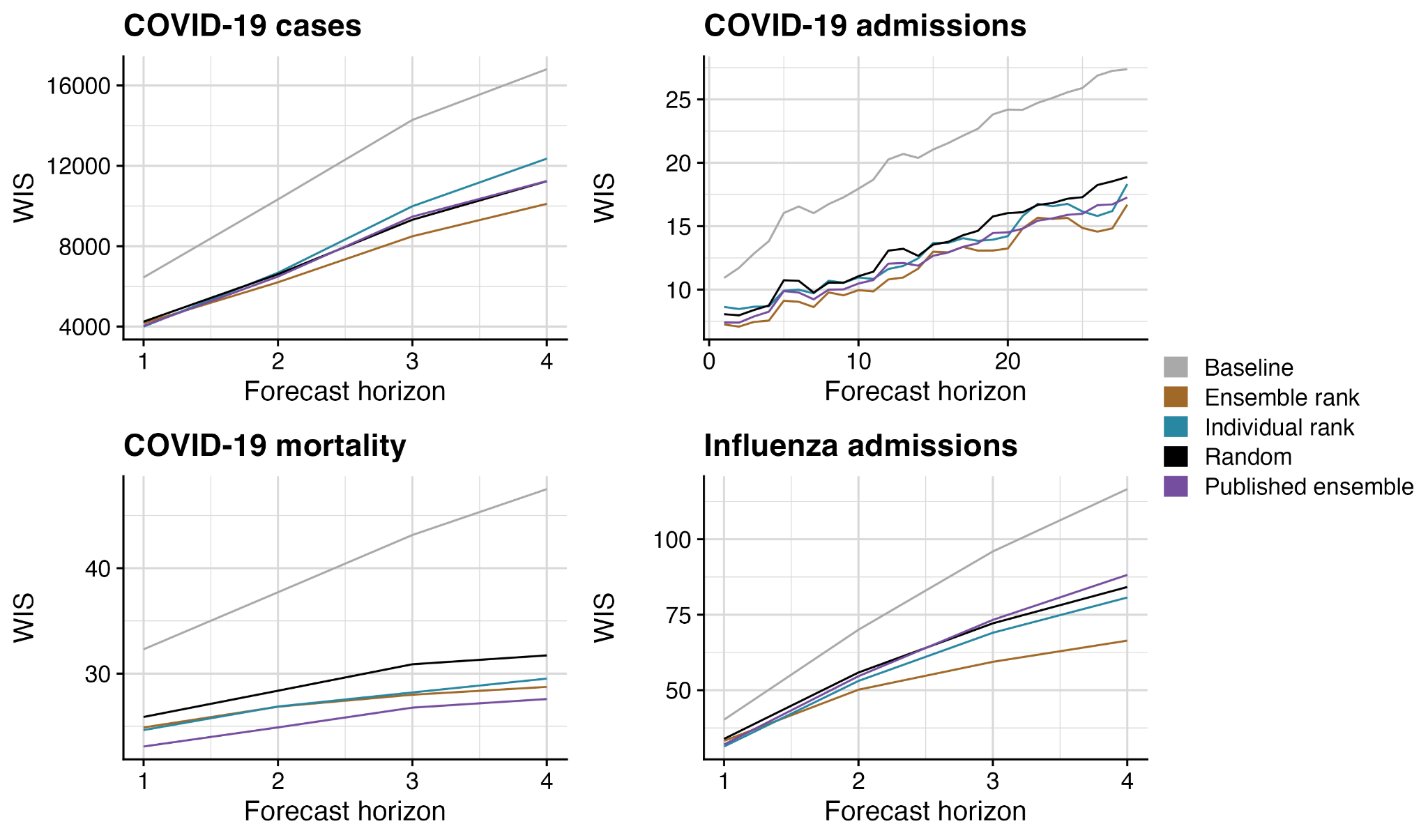

**Figure S14: Model performance for each forecasted target (horizon) across all recent collaborative hub efforts.** Average forecast score for each model and each target horizon across all forecast dates and locations from the testing period of the analysis. Lower scores indicate better forecast performance. Performance is visualized for ensembles of size four for the Ensemble rank, Individual rank, and Random ensembles, and only the mean performance is shown for the Random ensemble.

#### Location-specific forecast performance

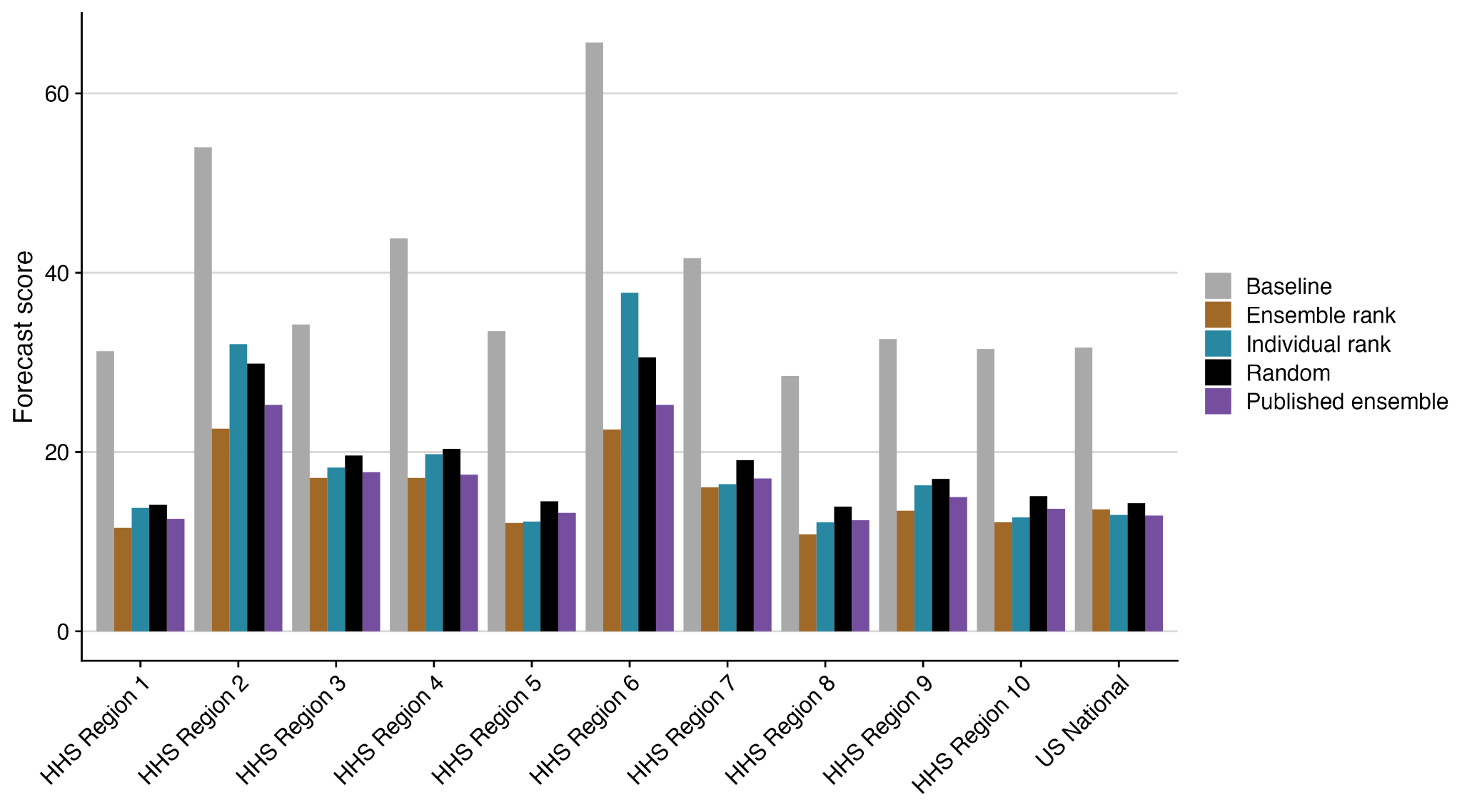

**Figure S15: Influenza-like illness (ILI %) model performance for each forecasted location.** Average forecast score for each model and location across all forecast targets and dates from the testing period of the analysis. Lower scores indicate better forecast performance. Performance is visualized for ensembles of size four for the Ensemble rank, Individual rank, and Random ensembles, and only the mean performance is shown for the Random ensemble.

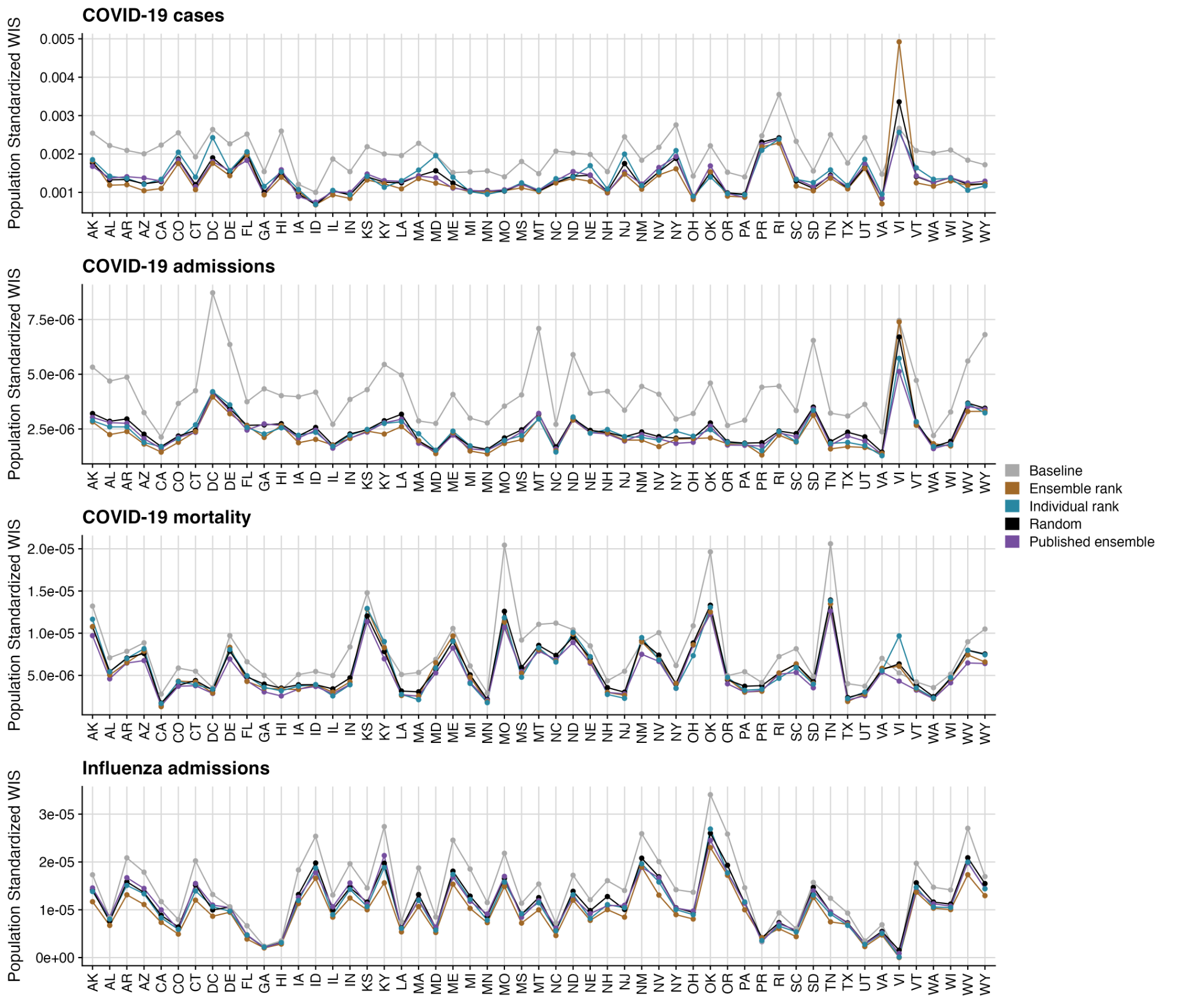

**Figure S16: Model performance for each forecasted location across recent collaborative hub efforts.** Average population standardized forecast score for each model and each location across all forecast dates and targets from the testing period of the analysis. WIS was divided by the region’s population to account for the absolute nature of the error metric. Lower scores indicate better forecast performance. Performance is visualized for ensembles of size four for the Ensemble rank, Individual rank, and Random ensembles, and only the mean performance is shown for the Random ensemble. Regions are ordered alphabetically by the abbreviation.
